# Weight loss with different types of fasting in patients with type two diabetes and hypertension: a randomized clinical trial

**DOI:** 10.1101/2024.10.10.24315274

**Authors:** Kuat Oshakbayev, Alisher Idrissov, Altay Nabiyev, Kenzhekyz Manekenova, Gulnara Bedelbayeva, Aigul Durmanova, Gani Kuttymuratov, Nurzhan Bikhanov, Bibazhar Dukenbayeva

## Abstract

**Background:** The coexistence of type two diabetes (T2D) and cardiovascular diseases (CVD) in patients with overweight increases the risk of macro-vascular complications. Some studies are controversial of the results of Ramadan fasting (RF) in patients T2D+CVD.

**Objective:** To compare different RF (antique and modern) on anthropometric, glycemic, blood pressure (BP), lipids, heel bone mineral density (HBMD), and ejection fraction (EF) in patients with T2D+CVD.

**Methods:** An open, 60-day, controlled, single-center, randomized clinical trial included 51 patients (29 women) with T2D+CVD: 26 in Main (antique RF); 25 in Controls (modern RF). *Primary endpoints:* weight loss, fasting blood glucose and, blood insulin, BP. *Secondary endpoints:* blood lipids, HBMD, and EF.

**Results:** Patients in Main lost weight –8.02 kg (*P*<0.0001), in Controls lost weight –2.67 kg (*P*<0.025); BMI in Main significantly decreased (*P*<0.0001), but in Controls did not significantly decrease (*P>*0.025). During a 30-day follow-up, Main did not regain weight and WC; but Controls regained weight and WC.

In Main BP, all glycemic parameters (fasting glucose, immunoassay insulin, HOMA-IR), lipids (cholesterol, triglyceride, HDL), HBMD, and EF in patients with and without heart failure (HF) were significantly normalized at 30-day RF (*P*<0.0001). In Controls BP and all glycemic parameters significantly improved but they did not achieve normal (*P*>0.025). In Controls HBMD and EF in patients with HF did not change (*P*>0.025). At 30-day follow-up, glycemic, BP, lipids, HBMD, and EF parameters did not significantly change in Main; but in Controls they worsened as weight regained.

**Conclusion:** Both the RF methods allowed weight loss, but antique RF led to markedly weight loss and significant positive change in glycemic, BP, lipids, bone mineralization, and EF in patients with T2D+CVD; the antique RF allowed patients to stop taking medications. The duration of the positive results depended on the maintenance of the achieved weight loss on RF.

**Trial Registration:** ClinicalTrials.gov NCT06410352 (05/08/2024): https://register.clinicaltrials.gov/prs/app/action/SelectProtocol?sid=S000EG8K&selectaction=Edit&uid=U0006MBT&ts=139&cx=bhsvd1

## 1 INTRODUCTION

Numerous studies show that the most common risk factor for developing type two diabetes mellitus (T2D), hypertension, heart failure (HF), and atherosclerosis is overweight/obesity. [1–4] T2D with cardiovascular diseases (CVD) are the leading cause of mortality/morbidity worldwide. [5, 6]

Hypertension and T2D are independently associated with impaired left ventricular diastolic function, independent of the effect of overweight/obesity and other covariates. [7, 8] HF is positively associated with overweight and T2D. [8, 9] The coexistence of hypertension and T2D increases the risk of macro-vascular complications and it is responsible for a high cardiovascular mortality. [10, 11] Early and aggressive treatment of the patients is therefore mandatory in diabetic and pre-diabetic groups. [12] Pharmacologic and biologic treatment of T2D and CVD has yielded positive results, however, these results were accompanied by numerous side effects. [5, 13, 14] Establishment of reliable, safe, and natural methods for the treatment of the non-communicable diseases, particularly in patients with T2D, hypertension, and HF, are required.

Several studies showed that weight loss improved left ventricular function in patients with elevated blood pressure (BP) and T2D, [1, 15, 16], and decreased mortality and morbidity from HF. [17–19] Despite adverse association between obesity and the non-communicable diseases, numerous studies have documented an obesity paradox, in which obese people with established CVD, including hypertension, coronary heart disease, and HF, have a better prognosis than patients who are not overweight or obese. [20–22] The cluster of T2D, CVD, dyslipidemia, and hypertension have also been associated with increased risk of low bone mineral density, [23, 24] together they may lead to macro-vascular catastrophe. [24, 25]

In countries where the official religion is Islam, there are large populations of Muslims with T2D and CVD who fast during Ramadan fasting (RF). RF lowers body weight, body fat percentage and body mass index (BMI). [26, 27] Changes in the pattern of meal and fasting during this long-fasting hours could lead to weight loss and improve glycemic and cardiovascular parameters. [26–28]

Some studies are controversial of the results of RF in patients T2D and CVD [26, 29], and it may cause a transient worsening of glycemic control that might be associated with weight gain [30, 31] and/or exacerbation of chronic diseases during RF. [32, 33] The majority of patients with stable cardiac illness can opt for RF safely. [26, 34] However, the effects of RF on cardiovascular outcomes and T2D remain uncertain. The relationship between weight reduction and improvement in health parameters in patients with T2D, serious CVD is not thoroughly studied.

Results of many studies suggest that the effect of RF on glycemic and/or cardiovascular parameters depends on the amount of weight loss during the RF. [26, 28, 35] We recently demonstrated weight loss with positive clinical results in overweight/obese patients with T2D and CVD. [36–38]

Ancient civilizations had another culture of RF than modern RF. [31, 39] For instance, antique fasting modes (Shabbat, Ramadan, Christian) prohibited eating meat and fatty foods. [39–41] It is assumed that modern RF differ from ancient RF. Antique people could lose more weight during RF than modern people. Ancient people did not have access to excess food consumption like modern people. The aim of this study was to compare the results of different RFs (ancient and modern) on the anthropometric, glycemic, BP, lipids, and ejection fraction (EF) in patients with T2D and CVD.

## 2 METHODS

### 2.1 Study Design

An open, 60-day, controlled, single-center, randomized clinical trial.

### 2.2 Participants

The study screened in total 96 participants, and in the intention-to-treat analysis enrolled for eligibility 61 adult treatment-seeking patients aged 35 to 65 years with T2D and hypertension; 35 were excluded due to inclusion criteria. From the enrolled 61 patients five patients were excluded due to noncompliance with the inclusion/exclusion criteria as explained below. Finally, 56 patients were included in the study. (**Figure 1**)

The patients were randomly divided into two groups: in Experimental group (Main) 28 patients; in Control group (Controls) 28 patients. Main group were on ancient (antique) RF, and Controls were on modern RF (conventional, traditional).

Five of the remaining 56 patients (7.1%) dropped out before study completion: two refused the RF for 2-3 days after starting, three were excluded due to noncompliance. Thus, 51 patients (29 women, 39 patients ≥50 years old, all Asian ethnicity) were included for the final analysis: in Main 26 patients; in Controls 25 patients. After 30-day of RF all patients were observed in 30-day followed-up. (**Figure 1**) In Main 14 patients and in Controls 13 patients had HF in their medical records and anamnesis with reduced EF <55%; I-II stages of the NYHA functional classification.

In total were 10 visits: Visit 1-2 (pre-baseline and baseline) – informed consent, blood samples, anthropometry & BP, ultrasound imaging & densitometry; Visits 3-5 (every week during RF) – medical evaluation & BP; Visit 6 (after the end of RF) – anthropometry & BP, blood samples, ultrasound & densitometry; Visits 7-9 (every week during 30-day of follow-up after RF) – medical evaluation & BP; Visit 10 (after follow-up period) – anthropometry & BP, blood samples, ultrasound & densitometry.

A combination of in-person conversations and telephone calls were conducted during the whole 60-day study period. During all visits, RF patients were closely followed and motivated (answers to questions, consultations, advice) by the clinical nutrition team to ensure weight loss in both the Main and Control groups.

#### Inclusion criteria

1) written informed consent; 2) 35≤ age <65 years old; 2) BMI ≥23 kg/m^2^; 3) diagnosis of T2D and hypertension ≥3 years and treatment with non-hormonal conventional pharmacological medications; 4) any level of triglycerides, cholesterol; 5) weight loss for 30-day RF, and 30-day of follow-up periods.

#### Exclusion criteria

1) age <35 or ≥65 years old; 1) insulin-depended T2D and uncontrolled high BP; 2) patients after bariatric surgery; 3) patients taking gastrointestinal lipase inhibitors, etc.; 4) presence of acute infection diseases or exacerbation of chronic diseases; 5) EF <40%; 6) history of alcohol consumption >30 g/day within the past 5 years; 7) malignancy within the past 5 years; kidney failure; gestation/lactation; hereditary diseases; or known hypersensitivity to any substances. ***Randomization.*** An independent statistician unconnected with clinical practice used simple randomization based on a single sequence of random assignments. A simple randomization method was tossing a coin. In the two treatment groups (Main vs. Controls), the side of the coin (heads/tails for Main/Controls as a random number generator) determines the assignment of each patients.

#### Outcome measures

*Primary endpoints:* weight loss, fasting blood glucose, blood insulin, systolic/diastolic BP (SBP/DBP). *Secondary endpoints:* blood lipids, heel bone mineral density (HBMD), and EF.

#### Interventions

Main group adhered to fasting on ancient RF; who strictly adhered to two principles of nutrition during ancient RF: 1) “no blood, no smell of fat” which means “don’t eat meat and don’t eat fatty foods” in the allotted time; 2) the ‘Sehri time’ (a meal eaten before dawn) included just 1.5-2.0 glass of water and fruits of 80-100 gr (dried apple, dates, and persimmon). After 30-day ancient RF, Main was followed by a 30-day time-restricted feeding (TRF) mode 20:4, wherein individuals fast for 20 hours with 4-hour window for caloric intake; or taking one meal a day (in the evening) with usual regular meal including meat and fatty foods. [42, 43]

Controls adhered to fasting on modern RF (conventional standard RF) with: 1) no restrictions to eat meat and/or fat meal in the allotted time; 2) no food restrictions to the ‘Sehri time’ and ‘Iftar time’ (a meal eaten after sunset). After a 30-day modern RF, Controls switched to a regular diet.

Both groups at baseline received conventional anti-diabetic, anti-hypertensive, pathogenetic, and symptomatic pharmacologic therapy. T2D was treated with Metformin, Sulfonylurea, GLP-1 agonists (exenatide, liraglutide), Glitazones in different combinations; hypertension was treated with beta-blockers, calcium channel blockers, diuretics, SGLT-2 Inhibitors (empagliflozin, canagliflozin) in different combinations. Both groups adhered to walking at least 3,000 steps/day, and sexual self-restraint.

#### Blinding of investigators

All patients were under the supervision of only four physicians from the clinical nutrition group; patients in both comparison groups were under the supervision of the same physicians without assignment to any patient group; different patients in the comparison groups were under the supervision of the same physician, and vice versa, the same patients in different comparison groups were under the supervision of different physicians.

### 2.3 Analytical Assessment

*Anthropometrical indicators* included age (years), weight (kg), BMI (kg/m^2^), WC (cm). Body composition parameters including fat mass (in % of total body weight), fat free mass, total body water, muscle mass, bone mass were measured using a Tanita-SC330S Body Composition Analyzer (Tanita Corp., Tokyo, Japan).

On the same blood samples, complete blood cell count, glucose, total cholesterol, high-density lipoprotein (HDL) and triglycerides were determined. Physical activity of patients assessed by the number of steps measured by individual pedometers in mobile phone or by Hoffmann-La Roche’s pedometers (Ltd.).

#### Hormonal assays

Fasting serum insulin (nU/L) was determined by immunoassay (Immunotech Insulin Irma kit, Prague, Czech Republic). Hyperinsulinemia was considered >12.5 nU/L. The Homeostasis Model Assessment insulin resistance indexes (HOMA-IR) was used as surrogate measure of insulin sensitivity as follows: HOMA-IR = ((fasting insulin in nU/L) × (fasting glucose in mmol/l)/22.5). Insulin resistance was considered if the index was > 2.

#### Imaging

Ultrasound imaging (GE Vivid 7 Ultrasound; GE Healthcare Worldwide USA, Michigan) used for echocardiographic examination, and HBMD measured using Lunar Achilles Express Ultrasound densitometry (GE Healthcare USA, Madison; at normal range 100.0±15.0 Units).

The patients temporarily stopped taking antidiabetic and antihypertensive medications 24 hours before BP measurement and blood collection to determine glucose and insulin at baseline. The standard of the American Diabetes Association (2017) was used for T2D diagnosis. [44] Hypertension was diagnosed by BP readings and from medical records. We carried out anthropometrical, laboratory, and instrumental examinations three times – before RF, after RF, and after a 30-day follow-up.

### 2.4 Statistics

#### Justification of the sample size

The estimated treatment difference will set to 10% with a standard deviation of 8% and the superiority margin of 5% (δ=0.05) [45] based on two-sided hypothesis testing. Using SPSS,Sample-Power,V23.0, the number of evaluable individuals needed per treatment arm ≥31. At least 96 patients we screened and recruited, and 61 patients were assessed for eligibility in the comparative clinical trial (**Figure 1**). [46] To increase in the power of the statistical analysis and receive significant results we used two-sided Student’s t tests with Bonferroni correction (*P-*values/2); where *P* values of <0.025 were set as significant differences in intra groups, and <0.025 between groups to compensate for the small number of patients (26 in Main vs 25 in Control). The study used SPSS Statistics v23 (SPSS Inc., Illinois, USA) and Microsoft Excel-2023 with normality assessed using histograms and box plots. Given the pilot nature of the study and firm hypotheses, were used. The study data is presented in Tables as Mean ± Standard Error of the Mean (M±SEM) for normally distributed data. All analyses were intention-to-treat.

## 3 RESULTS

### Anthropometric results (**Table 1**)

During the RF period patients in Main lost weight – 8.02 kg (*P*<0.0001), in Controls lost weight –2.67 kg (*P*<0.025); BMI in Main decreased significantly (*P*<0.0001), but in Controls BMI did not significantly decrease (*P>*0.025). Weight loss decreased due to fat mass in Main by –6.82% (*P*<0.0001) and in Controls by –1.38% (*P<*0.025); waist circumference in Main decreased by –9.36 cm (*P*<0.0001), but in Controls on –2.68 cm (*P<*0.025). In Main weight loss occurred due to abdominal component, and lean body mass did not significantly change in both groups.

**Table 1.**
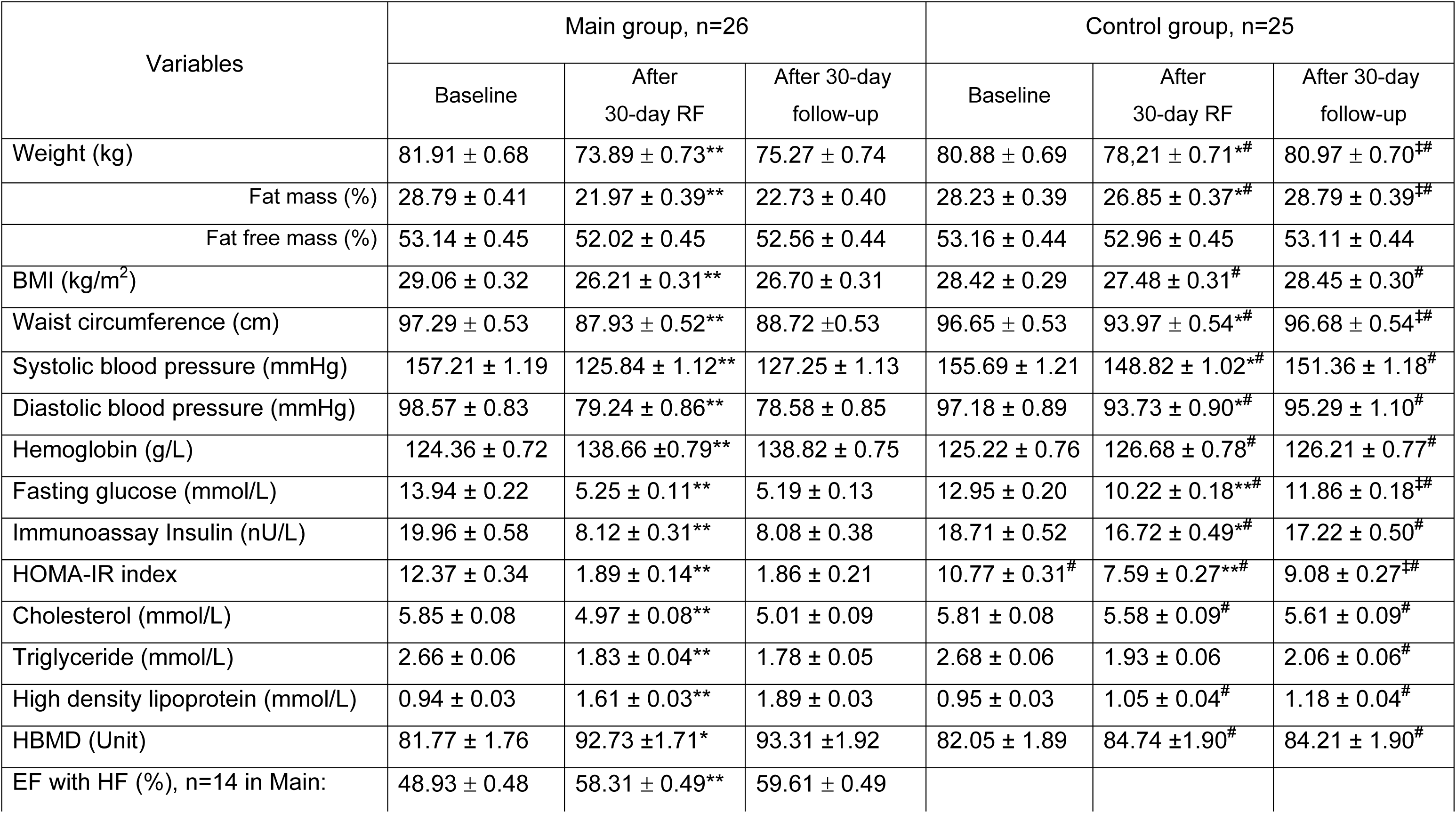
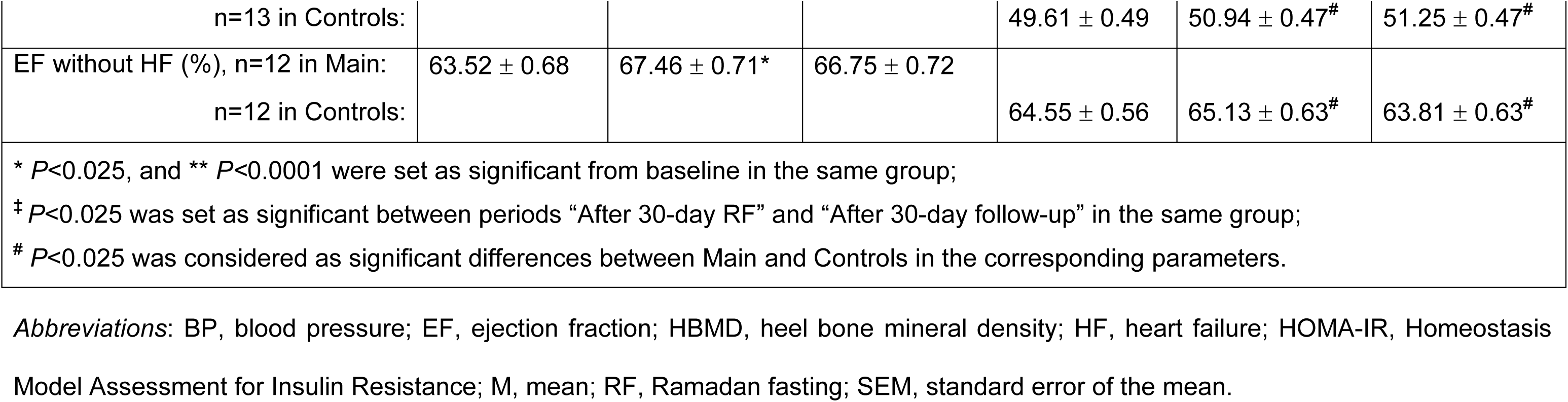
Anthropometrical data, body composition, blood pressure, glucose and lipid metabolism, HBMD, and EF in Main (n=26) and Controls (n=25) at baseline, after RF, and 30-day follow-up (M±SEM).

During a 30-day follow-up, Main tended to increase in weight and WC not significantly (*P=*0.19 and *P=*0.29, respectively); but Controls regained weight and increased in WC even more than at baseline significantly (*P=*0.010 and *P<*0.0015, respectively).

### Clinical and BP results (**Table 1**)

In Main SBP/DBP decreased starting from 5-7 days, and which significantly normalized at 30-day RF reaching recommended levels by the American Heart Association (2014) (*P<*0.0001); in Controls SBP/DBP significantly improved, but it did not achieve to normal ranges. After a 30-day follow-up, SBP/DBP remained within normal limits in Main, but they returned to previous ranges in Controls. After 3-5 days of the ancient RF, almost all patients in Main began to produce substantial amount of sputum, which occurred periodically throughout the entire RF period; this was not observed in Controls. Patients in Main complained less about thirst, healthy comfort, and clinical symptoms during RF compared to Controls, which may be due to abstinence from eating meat and fatty foods.

### Laboratory and instrumental (**Table 1**)

All glycemic parameters (fasting glucose, immunoassay insulin, HOMA-IR index) in Main significantly reduced (*P*<0.0001); in Controls the parameters also significantly decreased, but they did not reach normal. Lipids (cholesterol, triglyceride, HDL) significantly positive changed in Main (*P*<0.0001), but in Controls the parameters did not significantly change (*P*>0.025). At 30-day follow-up, glycemic parameters in Main did not significantly change; in Controls they worsened as weight regained.

Hemoglobin in Main significantly increased from baseline at a 30-day RF (*P*<0.0001), and the level was maintained at 30-day follow-up; in Controls it did not significantly change (*P*>0.025).

### *Imaging* (**Table 1**)

HBMD was significantly increased in Main (*P*<0.0001), in Controls it did not significantly change (*P*>0.025). At 30-day follow-up, HBMD in Main remained within normal limits.

EF in patients with HF and without HF significantly increased in Main after ancient RF (*P*<0.0001), and this achievement was maintained at 30-day follow-up. The EF parameters did not significantly increase in Controls neither after modern RF nor at 30-day follow-up (*P*>0.025).

Notably, in Main starting from 3-5 days of weight loss, it was necessary to reduce and completely stop taking the previous antidiabetic, antihypertensive, and other symptomatic medications in order to avoid hypoglycemic coma, hypotensive/hypotonic and other effects of the medications; by 6-10 days the drugs were stopped completely in Main, and at 30-day follow-up there was no evidence of T2D, hypertension, or relapse of HF. Patients in Controls could not reduce taking the previous medications, and they were receiving the same medication as at baseline.

## 4 DISCUSSION

T2D and CVD are the most common non-communicable diseases and they are problematic in countries with a higher rate of overweight. [4, 6, 10, 47] RF period lasted more than 20 hours during a 30-day RF (especially in summer), and the fasting period looked like the intermittent fasting practiced by millions of adult Muslims globally for a whole lunar month every year. [35, 48, 49]

In this open, 60-day, controlled, single-center, randomized clinical study among 51 patients with T2D in comorbidity with Hypertension, and HF (n=27), we found that significant improvements in glycemic profile, BP, lipids, EF, and HBMD were dependent on substantial weight loss. To our knowledge, this is one of the first studies to show that the different RFs may have variable treatment effects on patients with T2D and CVD. RF can have anti-hyperglycemic, antihypertensive, hypolipidemic, anti-osteoporotic effects, and restore EF, but this depends on the level of weight loss. The positive effects persisted if the weight did not return. Our study showed that when patients regained weight after a 30-day follow-up (Controls), their clinical results returned to baseline. They gained more weight and WC than baseline, and interestingly, fat mass tended to increase after the 30-day follow-up.

The TRF 20:4 mode allowed to maintain the achieved weight loss; they did not significantly gain weight at a 30-day follow-up and body structure also did not change. It is clinically important to maintain the achieved weight loss. Many studies evidence that TRF 20:4 allows to maintain the achieved weight loss [42, 50]

Obviously, great weight loss improved anabolic processes, because hemoglobin, HDL, HBMD levels significantly increased in Main. Abstinence from fatty foods and meat has an anti-osteoporosis effect and improves bone metabolism. [51, 52]

The study showed that patients in both groups lost weight due to a decrease in fat mass. The humans and animals studies indicated that temporal restriction diets did not induce muscle atrophy or proteolysis. [53, 54]

The Comprehensive Assessment of the Long-term Effects of Reducing Intake of Energy (CALERIE) phase 2 trial showed that a moderate reduction in energy intake could improve cardiometabolic health, and diabetic parameters in humans. [55, 56] Caloric restriction has been shown to extend the lifespan and health span of many animal species. [57] There has been growing interest in evaluating related strategies, such as intermittent fasting, periodic fasting, and TRF may achieve the univocal benefits of RF season. [43]

Our study also revealed that markedly weight loss on ancient RF allowed to withdraw symptomatic drugs due to necessary to reduce and completely stop taking previous medications. Reduction to complete discontinuation of previous prescribed symptomatic medications occurred as a result of great weight loss, and we had no intention of stopping the medications. Controls was unable to reduce previous medications due to insufficient weight loss.

Changes in diet and fasting during the ancient RF (long-fasting hours) resulted in noticeable weight loss and may have been a type of intermittent fasting. [26–28] According to our study results, the ancient RF was the same as intermittent fasting 20:4, because the patients had one meal a day. [29, 58] It was important that patients in Main strictly adhered to Ramadan’s principles (the ancient RF) as “don’t eat meat with don’t eat fatty foods”, and having only fruit meal on ‘Sehri time’.

RF requires abstinence from water and other drinks; it’s like dry fasting. Eating meat requires to drink more water, [59] so without meat patients in Main could handle the dry fasting. Indeed, Main group followed a diet in which they ate one meal a day, both during RF period and during a 30-day post-RF follow-up.

Overweight is a biological burden for the body, [60] and sexual self-restraint during RF also matters; because vigorous sex also reduces overall body energy and shortens life expectancy. [61–63] Most of the body metabolism is expended on the digestive and reproductive systems. [64]

Weight loss creates ‘body potential power to weight gain’ that can increase physical/mental activity, recovers from diseases, or weight regain. Individual body weight and individual limit of weight gain mode may explain the ‘obesity paradox’. [65, 66] The ‘Obesity paradox’ is driven by individual body weight and individual limit of weight gain mode. This will also lead to better and more personalized treatment for patients with T2D and CVD. The ability to accumulate fat mass is one of the foundations for survival in conditions of food shortage. But, the survival rule “eat any time, any place, any opportunity” often led to overweight. Therefore, periodic RFs allowed people to stay healthy and avoid obesity-related diseases for centuries. [67, 68]

### Limitations

The limitations of this study are: the study was a single-center study; this study was small and with a short follow-up duration. The strengths of this study are: Different types of RF have different effects on the health of patients with T2D and CVD and this depends on the degree of weight loss during it; both types of RF had positive effects on the patients in a randomized clinical trial; the positive results depended on maintaining the achieved weight loss during the RF. Future research should unpick the impact mechanisms of weight loss during Ramadan fasting on patients with T2D and CVD.

## 5 CONCLUSION

Both the RF methods allowed weight loss, but ancient RF led to markedly weight loss and significant positive change in glycemic, BP, lipids, bone mineralization, and EF in patients with T2D and CVD; the ancient RF allowed patients stopping medications. The duration of the positive results depends on the maintenance of the achieved weight loss; TRF 20:4 allows to maintain the achieved weight within 30-day follow-up.

## List of abbreviations

BMI: body mass index

BP: blood pressure

CVD: cardiovascular diseases

EF: ejection fraction

HbA1c: glycosylated hemoglobin A1c

HBMD: heel bone mineral density

HDL: high-density lipoprotein

HF: heart failure

HOMA-IR: Homeostasis Model Assessment for Insulin Resistance

M±SEM: Mean ± standard error of the mean

NCDs: noncommunicable chronic diseases

RF: Ramadan fasting

SBP/DBP: systolic/diastolic blood pressure

T2D: type two diabetes mellitus

TRF: time-restricted feeding

WC: waist circumference.

## Data Availability

The data are available from the authors upon reasonable request. Those wishing to request the study data should contact Principal Investigator of a research grant: Dr. Oshakbayev Kuat (, phone + 77013999394).

## ACKNOWLEDGMENTS

The authors thank the Diagnostic Center of University Medical Center and ANADETO Medical Center for recruiting patients, collecting data for the study, and providing technical assistance.

## Consent for publication

All the authors of the manuscript affirm that they had access to the study data and reviewed and approved the final version.

## Author contributions

*KO:* study design, data collection, diagnosis, and treatment of the patients, bibliography review, statistical analysis, data interpretation, writing of the paper with re-review. *AI:* study design, data collection, preparation of statistical data in Excel, diagnosis and treatment of the patients, bibliography, and paper review. *AN:* study design, introduction and part of methods writing, data interpretation, and paper review. *KM:* data collection, data interpretation, bibliography, statistical analysis, discussion writing, and paper review. *GB:* study design, results and discussion writing, and paper review. *AD* and *NB*: data collection, data interpretation, results and discussion writing, and paper review. *BD:* study design, data collection, preparation of the statistical data in Excel, data interpretation, discussion writing, bibliography, and writing the paper with review. All the authors read and approved the final manuscript.

## Declaration of Generative AI and AI-assisted technologies in the writing process

During the preparation of this work the authors did not use it.

## Figure legends

Figure 1. Participant CONSORT Flow Diagram The Ramadan fasting weight loss in patients with type two diabetes, Hypertension, and with Heart failure: a randomized clinical trial

